# Frequent shedding of multi-drug resistant pneumococci among adults living with HIV on suppressive antiretroviral therapy in Malawi

**DOI:** 10.1101/2022.10.28.22281638

**Authors:** Lusako Sibale, Joseph Phiri, Ndaona Mitole, Newton Kalata, Tinashe Nyazika, Alice Kalirani, Mercy Khwiya, Gift Sagawa, Deus Thindwa, Todd D Swarthout, Neil French, Ken Malisita, Arox Kamng’ona, Daniela M Ferreira, Robert S. Heyderman, Brenda A. Kwambana-Adams, Kondwani Jambo

## Abstract

People living with human immunodeficiency virus (PLHIV) on antiretroviral therapy (ART) are reported to have three times higher carriage of *Streptococcus pneumoniae* than their HIV-uninfected counterparts in point prevalence studies. Using a longitudinal cohort study design, we assessed pneumococcal carriage density, shedding and antibiotic resistance profiles, as well as nasal mucosal immunity, in otherwise healthy PLHIV on ART for at least one year, compared to HIV-uninfected participants in Malawi. Pneumococcal carriage density was higher among PLHIV compared to HIV-uninfected participants. Moreover, PLHIV were twice more likely to shed pneumococci than HIV-uninfected participants. In PLHIV, aerosol shed pneumococci were more often multi-drug resistant (MDR) than nasopharyngeal carried isolates recovered from the same individual. Consistent with high shedding, PLHIV exhibited heightened neutrophil-mediated nasal mucosa inflammation. We propose that PLHIV should be considered in intervention strategies, such as vaccination, as they could be an important reservoir for transmission of MDR *S. pneumoniae*.

## Introduction

*Streptococcus pneumoniae* has at least 100 distinct serotypes ^1, 2^ and is a common coloniser of the human nasopharynx. Pneumococcal carriage is a prerequisite for life-threatening invasive pneumococcal disease (IPD) including pneumonia, meningitis, and bacteraemia ^3^, and is critical for transmission ^4^. In high- and middle-income countries, infant pneumococcal conjugate vaccine (PCV) programmes have resulted in a significant reduction in pneumococcal carriage and IPD in both vaccinated children (direct protection) and unvaccinated older children and adults (indirect protection) ^5, 6, 7, 8^. However, in low-income countries such as Malawi, where the 13-valent PCV (PCV13) was introduced into the infant immunisation programme in 2011, there is limited indirect protection among unvaccinated older children and adults despite evidence of substantial direct protection against IPD among vaccinated children ^9^.

Moreover, pneumococcal carriage prevalence has been reported to be over two-fold higher in PLHIV on antiretroviral therapy (ART) (40-60%) than in those HIV-uninfected (8-15%) or ART-naïve PLHIV (18-25%) ^10, 11^. Residual carriage of all PCV13 serotypes (VTs) among Malawian adults living with HIV on ART has remained relatively high ^7, 11^, with low socioeconomic status having been shown to exacerbate overall carriage prevalence^12^.

Emergence of pneumococcal antimicrobial resistance (AMR) is associated with carriage and antibiotic usage ^13, 14, 15^. *S. pneumoniae* is among the priority bacterial pathogens due to its increased resistance to penicillin according to World Health Organisation (WHO) Global Antimicrobial Resistance and Use Surveillance System (GLASS) ^16^. It has been shown that PLHIV are more likely to take antibiotics, including cotrimoxazole prophylaxis to prevent opportunistic bacterial infections^17, 18^. Moreover, PLHIV are also at increased risk of disease caused by respiratory viruses compared to the general population^19, 20^. Common viral co-infections ^21, 22, 23^, including Respiratory Syncytial Virus (RSV) and Influenza ^24^, have been shown to promote high pneumococcal carriage density that in turn drives increased bacterial shedding ^21, 25, 26^, a surrogate for transmission potential ^27, 28^. Furthermore, high pneumococcal carriage density has been shown to associate with invasive pneumococcal disease ^29, 30^. Together, this may suggest that in addition to being at increased risk of invasive pneumococcal disease, PLHIV could be an underappreciated reservoir driving transmission of antimicrobial resistant pneumococci in the community, calling for an in-depth evaluation of this population.

Using a longitudinal cohort study, we investigated pneumococcal carriage density and shedding, as well as nasal immunity, in otherwise healthy PLHIV on ART for more than one year and HIV-uninfected participants. We show that PLHIV exhibit higher density pneumococcal carriage and more commonly shed antimicrobial resistant pneumococci. These findings have broader implications in the development of interventions to curb AMR pneumococcal disease and transmission.

## Results

### Clinical and demographic characteristics

Between July 2019 and August 2021, we screened 512 adults among whom 28% (144/512) had confirmed pneumococcal carriage and thus eligible for recruitment (Fig. 1). Of these, 37.5% (54/144) were HIV-uninfected and 62.5% (90/144) were PLHIV on ART for more than 1 year (median 5.5 years [IQR 2.8-10.1]) (Table S1). Due to Coronavirus disease (COVID-19) pandemic restrictions, 37.5% (54/144) did not complete the study follow-up period of 5 months (Fig. 1). Therefore, the final analysis is restricted to the 35 HIV-uninfected participants and 55 PLHIV that completed the 5 months follow-up period. PLHIV, compared to HIV-uninfected participants were older, with 44% (24/55) and 20% (7/35) >36 years old, respectively (p=0.049). Likewise, compared to HIV-uninfected participants, PLHIV had a lower socioeconomic status as measured by asset ownership index ^12^ (median 4 [IQR 2.0-5.0] vs. 6 [3.5-7.50], p=0.005), and a lower CD4 count (median 514 cells/μl [IQR 652-927] vs. 760 cells/μl [IQR [332-750], p<0.001) (Table 1).

**Figure 1.**
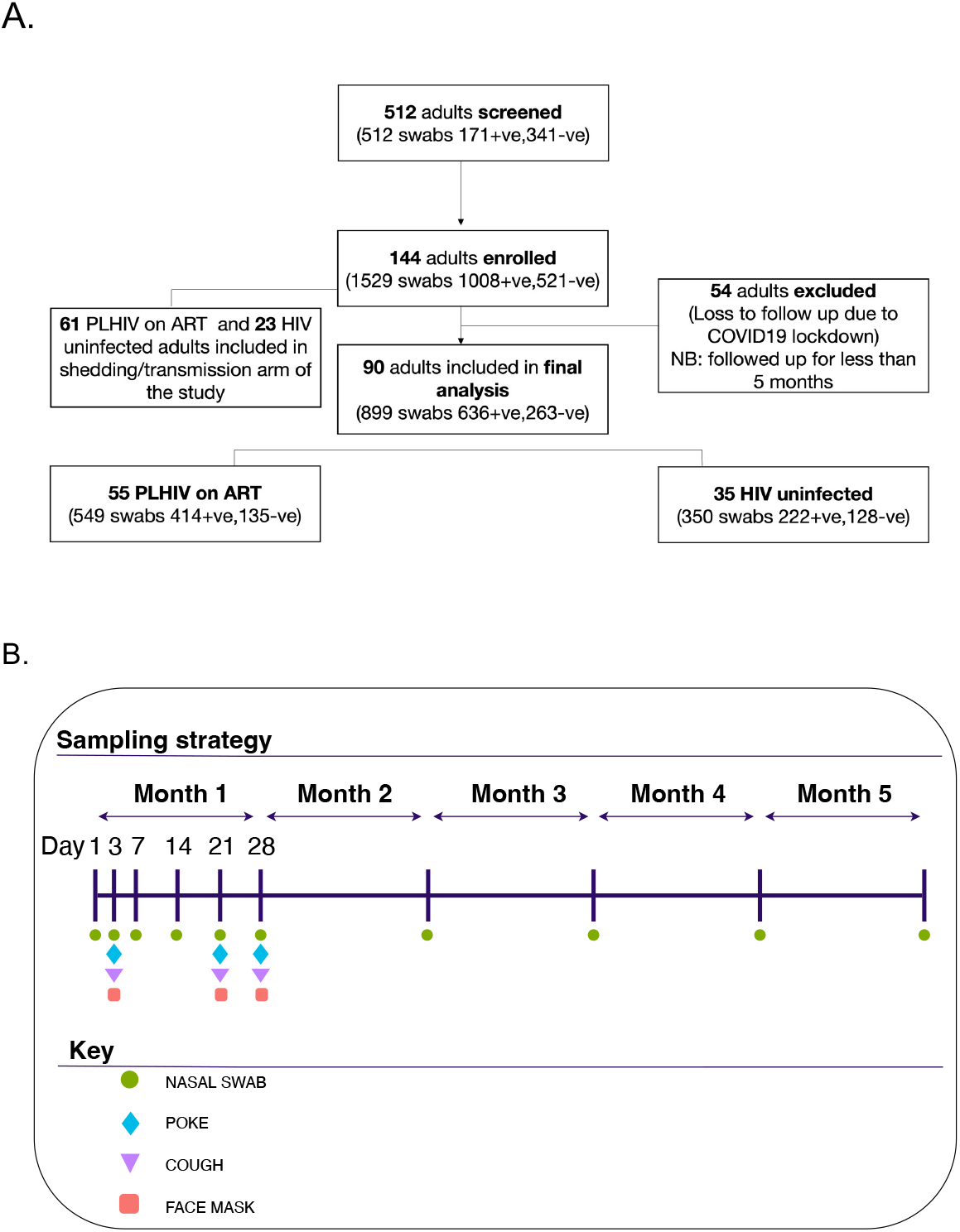
Recruitment flow diagram and study design. A) Flow diagram showing the number of adults and pneumococcal samples included in the analysis among PLHIV on ART and HIV uninfected participants. B) Sampling strategy, showing longitudinal follow up of study participants and sample collection points.

**Table 1.** Baseline characteristics

| Table 1. Baseline characteristics |  |  |  |
| --- | --- | --- | --- |
| Characteristic | HIV-, N = 35<sup>1</sup> | PLHIV, N = 55<sup>1</sup> | p-value<sup>2</sup> |
| **Sex** |  |  | 0.7 |
| Female | 25 (71%) | 41 (75%) |  |
| Male | 10 (29%) | 14 (25%) |  |
| **Age group** |  |  | **0.049** |
| 18-25 | 12 (34%) | 10 (18%) |  |
| 26-35 | 16 (46%) | 21 (38%) |  |
| 36-45 | 7 (20%) | 24 (44%) |  |
| **Number of U5 children per household** |  |  | 0.7 |
| 1 | 26 (74%) | 45 (82%) |  |
| 2 | 8 (23%) | 9 (16%) |  |
| 3 | 1 (2.9%) | 1 (1.8%) |  |
| **Socioeconomic status<sup>3</sup>** |  |  | **0.005** |
| Median (IQR) | 6.00 (3.50, 7.50) | 4.00 (2.00, 5.00) |  |
| **CD4 count (Cells/μl)** |  |  | **<0.001** |
| Median (IQR) | 760 (652, 927) | 514 (332, 750) |  |
| **ART duration (Years)** |  |  |  |
| Median (IQR) | NA (NA, NA) | 5.8 (3.4, 11.3) |  |
| **HIV viral load (Copies/ml)<sup>4</sup>** |  |  |  |
| Median (IQR) | NA (NA, NA) | 39 (39, 18,928) |  |

<sup>1</sup> n (%)
<sup>2</sup> Pearson's Chi-squared test; Fisher's exact test; Wilcoxon rank sum test
<sup>3</sup> Socioeconomic status score based a possession index which is calculated as a sum of positive responses for household ownership of each of the fifteen different functioning items such as watch, radio, bank account, iron (charcoal), sewing machine (electric), mobile phone, CD player, fan (electric), bednet, mattress, bed, bicycle, motorcycle, car, and television
<sup>4</sup> i.e. Only 10 PLHIV on ART>1yr had a detectable Viral load

### Higher NVT carriage positivity in PLHIV than HIV-uninfected participants

Overall (VT + non-VT [NVT]) pernasal swab pneumococcal positivity was higher among PLHIV compared to HIV-uninfected participants (72% [95% CI 68.2–76.2] vs. 59% [53.7–64.8] p<0.001) (Fig. 2a). Furthermore, NVT pernasal swab pneumococcal positivity was higher in PLHIV (55.5% [51.2–59.7] vs. 44.9% [39.6–50.2], p=0.002) (Fig. 2b). In contrast, VT pernasal swab pneumococcal positivity was similar between PLHIV and HIV-uninfected participants: 19.6% [16.4–23.2] and 18.6% [14.6-23.0], p=0.5777 (Fig. 2a). The dominant VT carriage isolates among both PLHIV, and HIV-uninfected participants were serotype 3 (49.1% and 43.1%) and serotype 19F (19.4% and 29.2%) respectively (Fig. 2c). Using a multivariable logistic regression model (adjusting for age, gender, season, socioeconomic status and HIV status), showed that PLHIV were more likely to carry NVT than HIV-uninfected participants (adjusted odds ratio (aOR) 1.4, (95% CI 1.1-1.9, p=0.015)) (Table S2a). In contrast, the likelihood of having VT carriage was similar between PLHIV and HIV-uninfected participants (aOR 1.0, (95% CI 0.7-1.5, p>0.9) (Table S2b). Together, these data show that high pneumococcal carriage prevalence in PLHIV is mostly driven by NVT, but also highlight high residual VT carriage of serotype 3 and 19F in both PLHIV and HIV-uninfected participants.

**Figure 2.**
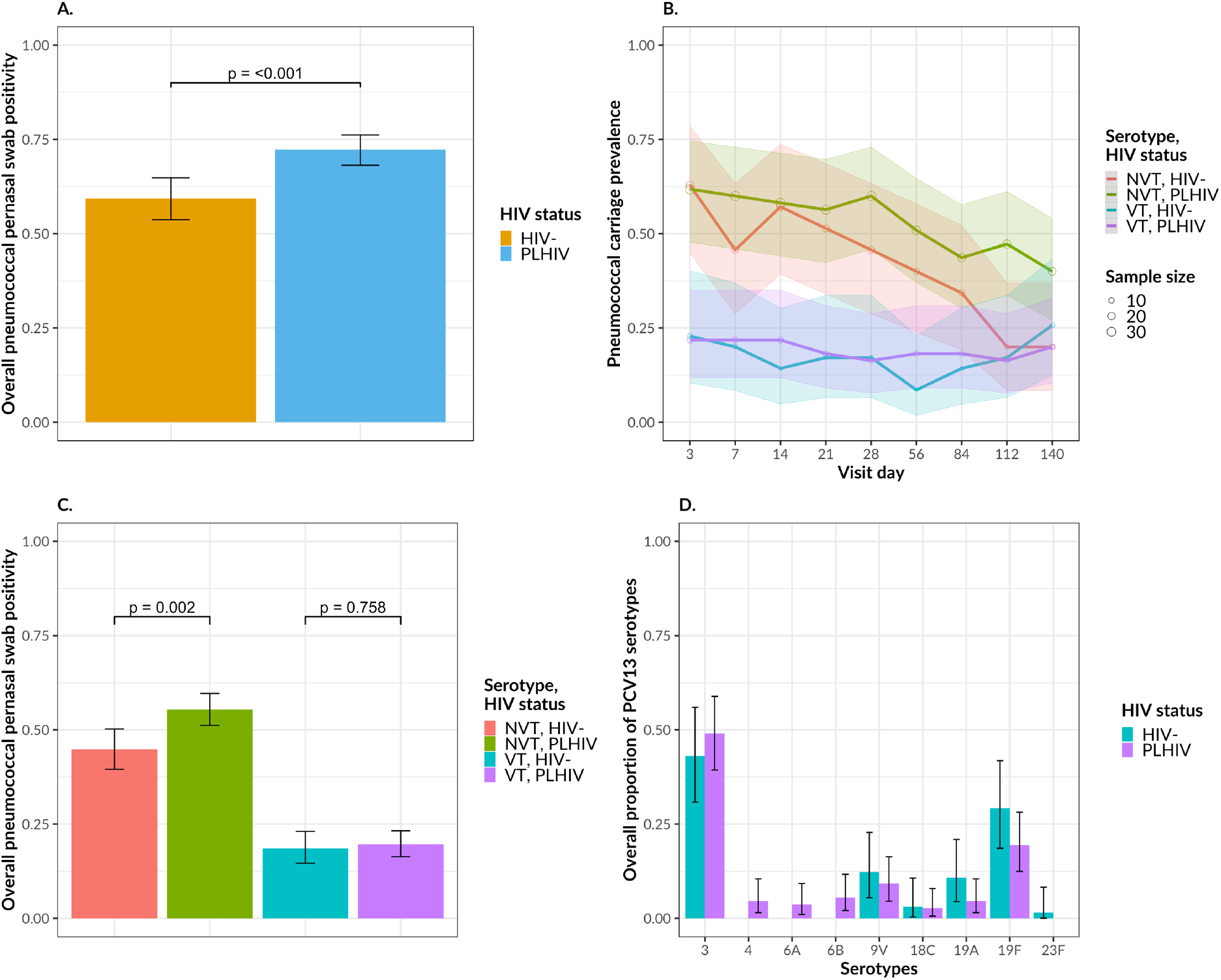
Overall (VT+NVT) pneumococcal swab positivity among PLHIV and HIV-uninfected adults. A) HIV-stratified pneumococcal swab positivity. B) HIV and serogroup stratified pneumococcal swab positivity by different nasopharyngeal sampling visits. C) HIV and serogroup stratified pneumococcal swab positivity. D) HIV-stratified proportion of PCV13 serotypes. The denominator for each serotype is the total PCV13 serotypes in each group. Black vertical lines represent 95% confidence intervals. Data were analysed using Chi-square test (HIV-uninfected: n=35, PLHIV: n=55). Source data are provided as a Source Data file. PCV13, pneumococcal conjugated vaccine 13; ART, antiretroviral therapy; NVT non-PCV13 serotype; VT, PCV13 serotypes

### Higher density pneumococcal carriage in PLHIV than HIV-uninfected participants

The overall median pneumococcal carriage density was higher among PLHIV (40,738.03CFU/ml [95% CI 16218.1-104712.9] vs. 10,000.00CFU/ml [4466.84-22908.68], p<0.001) (Fig. 3). In a multivariable logistic regression model (adjusting for sex, age, carriage density, season, and socioeconomic status), PLHIV were more likely to harbour higher density carriage than HIV-uninfected participants (aOR 1.6, 1.1-2.5, p=0.027) (Table 2). Collectively, these findings show high propensity for high-density carriage in PLHIV.

**Table 2.**
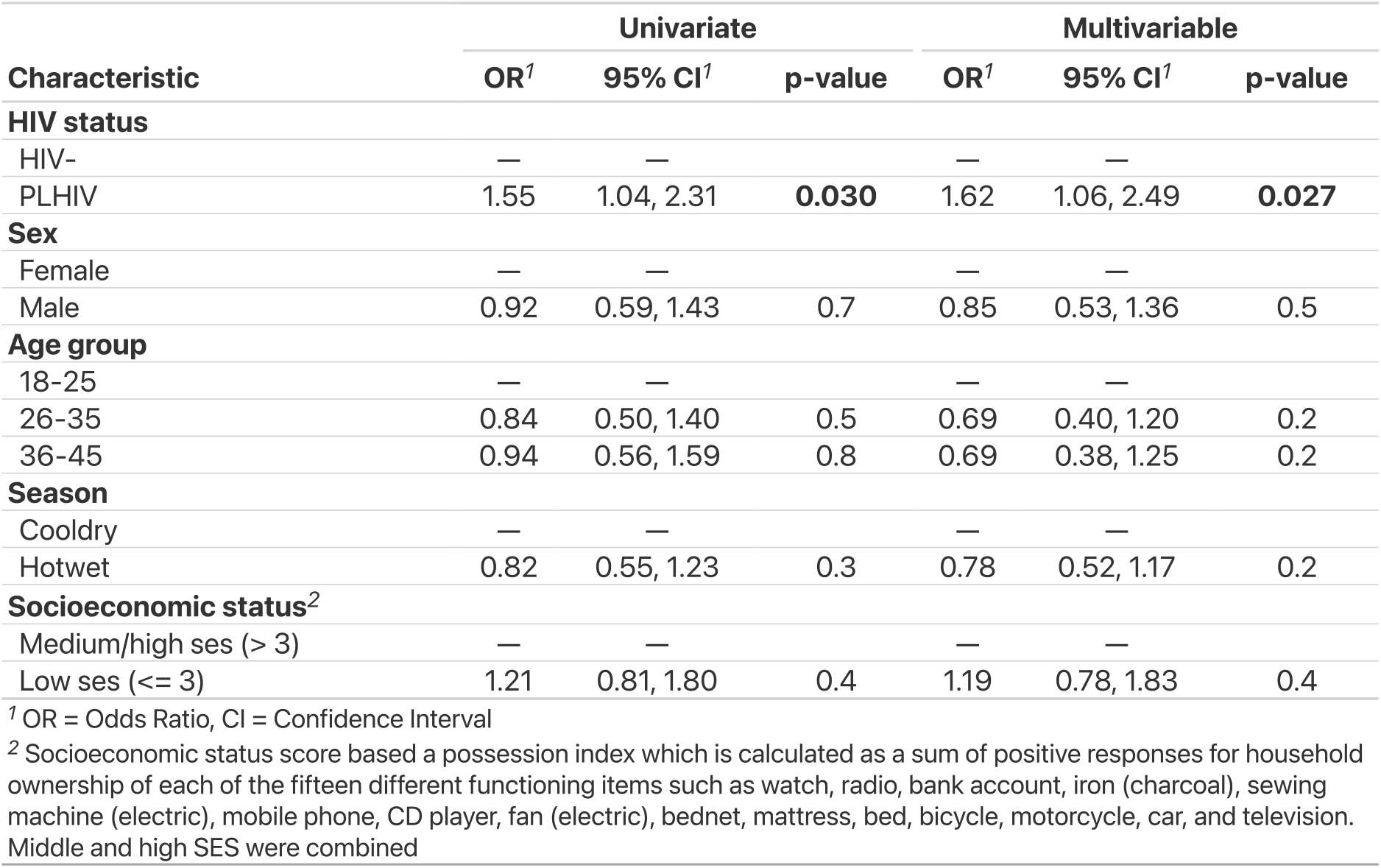
Pneumococcal density <=2010CFU/ml vs Pneumococcal density >2010FU/ml

| Table 2. Pneumococcal density ≤2010CFU/ml vs Pneumococcal density >2010CFU/ml |  |  |  |  |  |  |
| --- | --- | --- | --- | --- | --- | --- |
| Characteristic | Univariate |  |  | Multivariable |  |  |
|  | OR<sup>1</sup> | 95% CI<sup>1</sup> | p-value | OR<sup>1</sup> | 95% CI<sup>1</sup> | p-value |
| **HIV status** |  |  |  |  |  |  |
| HIV- | — | — |  | — | — |  |
| PLHIV | 1.55 | 1.04, 2.31 | **0.030** | 1.62 | 1.06, 2.49 | **0.027** |
| **Sex** |  |  |  |  |  |  |
| Female | — | — |  | — | — |  |
| Male | 0.92 | 0.59, 1.43 | 0.7 | 0.85 | 0.53, 1.36 | 0.5 |
| **Age group** |  |  |  |  |  |  |
| 18-25 | — | — |  | — | — |  |
| 26-35 | 0.84 | 0.50, 1.40 | 0.5 | 0.69 | 0.40, 1.20 | 0.2 |
| 36-45 | 0.94 | 0.56, 1.59 | 0.8 | 0.69 | 0.38, 1.25 | 0.2 |
| **Season** |  |  |  |  |  |  |
| Cooldry | — | — |  | — | — |  |
| Hotwet | 0.82 | 0.55, 1.23 | 0.3 | 0.78 | 0.52, 1.17 | 0.2 |
| **Socioeconomic status<sup>2</sup>** |  |  |  |  |  |  |
| Medium/high ses (> 3) | — | — |  | — | — |  |
| Low ses (≤ 3) | 1.21 | 0.81, 1.80 | 0.4 | 1.19 | 0.78, 1.83 | 0.4 |

<sup>1</sup> OR = Odds Ratio, CI = Confidence Interval
<sup>2</sup> Socioeconomic status score based a possession index which is calculated as a sum of positive responses for household ownership of each of the fifteen different functioning items such as watch, radio, bank account, iron (charcoal), sewing machine (electric), mobile phone, CD player, fan (electric), bednet, mattress, bed, bicycle, motorcycle, car, and television. Middle and high SES were combined

**Figure 3.**
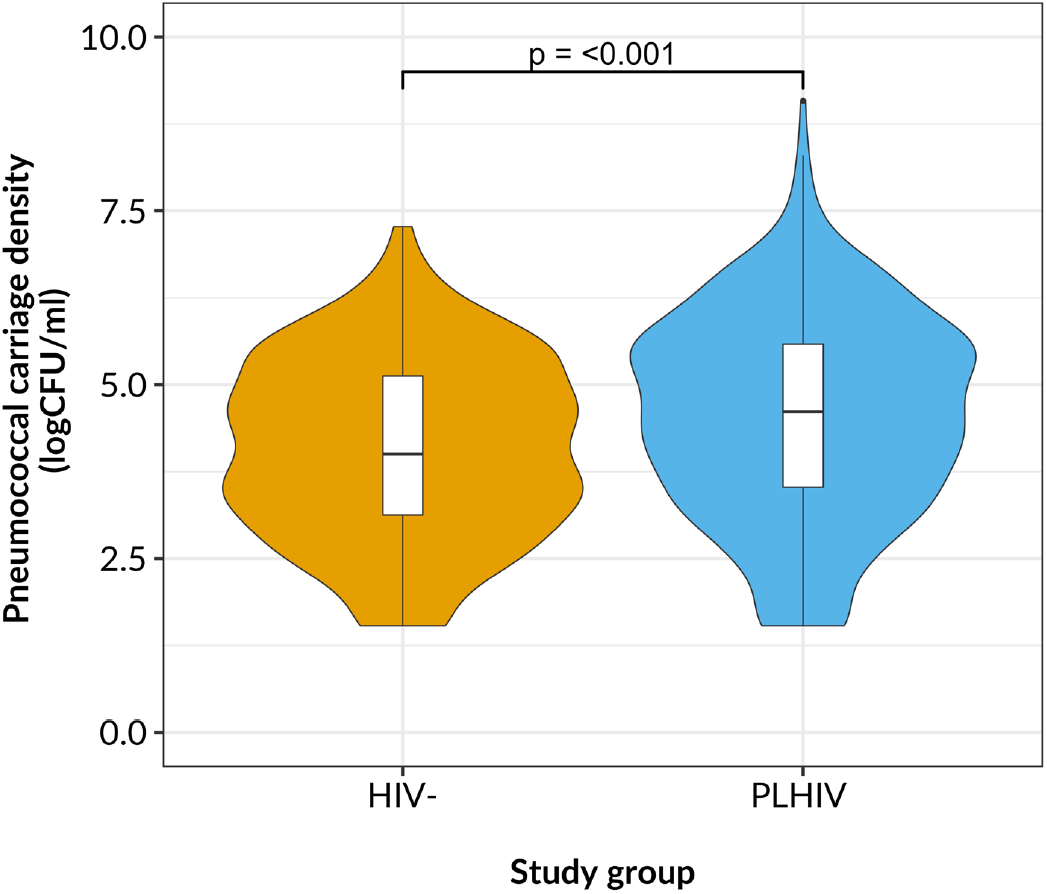
Pneumococcal carriage density among PLHIV and HIV-uninfected participants. Log median carriage density stratified by HIV status for 5 months study follow-up time. For all boxplots, box boundaries correspond to 25^th^ and 75^th^ percentiles; whiskers extend to a maximum of 1.5x the inter-quartile range, with values outside the box and whiskers being outliers. Data were analysed using Wilcoxon test (HIV-uninfected: n=35, PLHIV: n=55). Source data are provided as a Source Data file. ART, antiretroviral therapy; CFU, colony forming units

### Higher proportion of pneumococcal shedding in PLHIV than HIV-uninfected participants

Considering the high-density carriage in PLHIV, we sought to determine whether they were also more likely to shed pneumococci than HIV-uninfected participants. The overall proportion of adults shedding pneumococci using either sampling technique was significantly higher among PLHIV than HIV-uninfected participants (57.6% [95% CI 48.4-66.4] vs. 27.3% [18.3-45.4], p<0.001) (Fig. 3a). Further analysis revealed that NVT were shed more frequently in PLHIV than HIV-uninfected participants (31.6% [25.9-37.8] vs. 11.2% [5.7-19.2], p<0.001) (Fig. 3b). However, VT shedding was similar between PLHIV on ART and HIV-uninfected participants (6% [3.4-9.7] vs. 5.1% [1.7-11.5], p=0.946) (Fig. 3b). Among the shed VT isolates, serotype 3 (73.3% and 40%) and serotype 19F (13.3% and 60%) were dominant among both the PLHIV and HIV-uninfected participants, respectively (Fig. 3c). Moreover, the overall proportion of adults shedding pneumococci were significantly higher in both the mechanical (40.3% [95% CI 31.6-49.5] vs. 18.4% [8.8-32], p=0.010) and aerosol (36% [95% CI 27.6-45.1] vs. 14.3% [95% CI 5.9-27.2], p=0.009) samples in PLHIV than HIV-uninfected participants (Fig 3d-e).

To identify factors influencing pneumococcal shedding, we performed a multivariable logistic regression analysis that included age, gender, socioeconomic status, season, pneumococcal carriage density, and HIV status. HIV status and pneumococcal carriage density were significantly associated with pneumococcal shedding (Table 3). Specifically, PLHIV were twice more likely to shed any pneumococci than HIV-uninfected participants (aOR 2.4, 95% CI 1.04-5.7, p=0.039), while individuals with high-density pneumococcal carriage, irrespective of HIV status, were greater than three times more likely to shed any pneumococci than those with low-density carriage (aOR 3.37, 1.56-7.29, p=0.002) (Table 3). Together, these findings show that high proclivity for pneumococcal shedding in PLHIV, and in individuals with high density carriage (irrespective of HIV status).

**Table 3.**
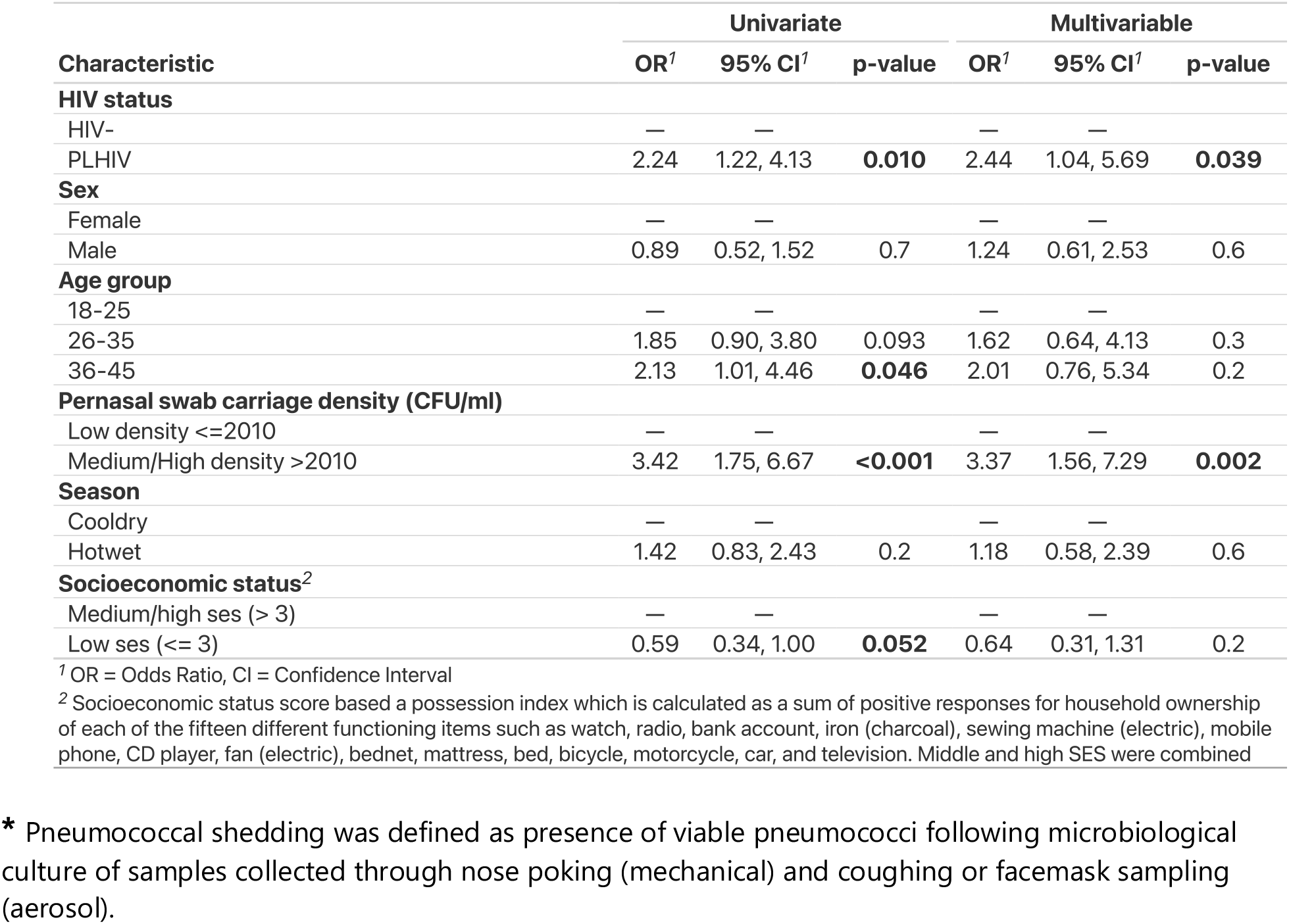
Factors associated with pneumococcal shedding

### PLHIV shed antimicrobial resistant pneumococci

Next, we sought to assess the antimicrobial susceptibility profile of the carried and shed isolates from PLHIV and HIV-uninfected participants. Overall, the antimicrobial susceptibility profile of nasopharyngeal carriage isolates was similar between PLHIV compared and HIV-uninfected participants (Table 5). However, the proportion of co-trimoxazole resistant isolates was significantly higher in PLHIV (98% [95% CI 88%-100%] vs. 76% [58%-89%], p=0.002) (Table 4). The overall proportion of non-susceptible tetracycline (Tet), benzylpenicillin (PenG), as well as multi-drug resistant isolates were significantly higher in aerosol shed isolates compared to nasopharyngeal carriage isolates in PLHIV for samples collected from the same individual (Tet, 61% [95% CI 46%-75%] vs. 26% [14%-41%], p<0.001; PenG, 61% [46%-75%] vs. 32% [19%-48%], p=0.010; MDR, 64% [48%-77%] vs. 30% [CI 17%-45%], p=0.001) (Table 5). In contrast, the AMR profile of mechanical shed isolates were similar to the nasopharyngeal carriage isolates (Table 6). Among HIV-uninfected participants, no differences were observed in the antimicrobial susceptibility profile between shed and carriage isolates, irrespective of the shedding route (Table S3). Together, these data show that PLHIV shed multi-drug resistant pneumococci, especially via the aerosol route.

**Table 4.** Baseline pneumococcal carriage isolates antibiogram

| Characteristic | HIV-, N = 35<sup>1</sup> | PLHIV, N = 57<sup>1</sup> | p-value<sup>2</sup> |
| --- | --- | --- | --- |
| **Cotrimoxazole** |  |  | **0.002** |
| Susceptible | 8 (24%) | 1 (2.0%) |  |
| non-susceptible | 26 (76%) | 50 (98%) |  |
| **Oxacillin** |  |  | 0.5 |
| Susceptible | 15 (43%) | 26 (51%) |  |
| non-susceptible | 20 (57%) | 25 (49%) |  |
| **Tetracycline** |  |  | 0.2 |
| Susceptible | 16 (47%) | 32 (63%) |  |
| non-susceptible | 18 (53%) | 19 (37%) |  |
| **Erythromycin** |  |  | **0.058** |
| Susceptible | 21 (62%) | 41 (80%) |  |
| non-susceptible | 13 (38%) | 10 (20%) |  |
| **MDR<sup>3</sup>** |  |  | 0.5 |
| non MDR | 26 (76%) | 42 (82%) |  |
| MDR<sup>3</sup> | 8 (24%) | 9 (18%) |  |

<sup>1</sup> n (%)<sup>2</sup> Fisher's exact test; Pearson's Chi-squared test<sup>3</sup> i.e. Oxacillin is excluded from MDR definition

**Table 5.** Pneumococcal shedding isolate antibiogram among PLHIV

| Table 5. Pneumococcal shedding isolates antibiogram among PLHIV |  |  |  |  |  |  |
| --- | --- | --- | --- | --- | --- | --- |
| Characteristic | Aerosol shedding vs<br>Nasopharyngeal carriage |  |  | Mechanical shedding vs<br>Nasopharyngeal carriage |  |  |
|  | Aerosol<br>shedding,<br>N = 44<sup>1</sup> | Nasopharyngeal<br>carriage, N = 44<sup>1</sup> | p-value<sup>2</sup> | Mechanical<br>shedding,<br>N = 50<sup>1</sup> | Nasopharyngeal<br>carriage, N = 50<sup>1</sup> | p-value |
| **Tetracycline** | 27 (61%) | 11 (26%) | **<0.001** | 16 (32%) | 15 (30%) | 0.8 |
| **Erythromycin** | 19 (43%) | 12 (28%) | 0.14 | 14 (28%) | 16 (32%) | 0.7 |
| **Cotrimoxazole** | 41 (93%) | 41 (95%) | >0.9 | 47 (94%) | 46 (92%) | >0.9 |
| **Benzylpenicillin<sup>3</sup>** | 27 (61%) | 14 (32%) | **0.010** | 19 (38%) | 14 (28%) | 0.4 |
| **MDR** | 28 (64%) | 13 (30%) | **0.001** | 15 (30%) | 15 (30%) | >0.9 |

<sup>1</sup> n (%)
<sup>2</sup> Pearson's Chi-squared test; Fisher's exact test
<sup>3</sup> i.e. Minimum inhibitory concentration

### PLHIV exhibit neutrophil-mediated inflammation in the nasal mucosa

Lastly, we sought to determine whether PLHIV have a more inflamed nasal mucosa than HIV-uninfected adults, as inflammation has been shown to play a significant role in pneumococcal shedding ^31, 32, 33^. To do this, we recruited PLHIV on ART for more than 1 year, and age- and gender-matched HIV-uninfected adult controls, irrespective of pneumococcal carriage status, from whom nasal cells and nasal lining fluid samples were collected weekly for 5 weeks (Fig. S1). Using flow cytometry-based immunophenotyping, we characterised the immune cell composition in the nasal mucosa, a representative gating strategy is shown in Fig 5a. The abundance of CD14^+^ monocytes (median ratio 0.01 [95% CI 0.001-0.011] vs 0.01 [95% CI 0-0.014], p=0.55) and CD3^+^ T cells (median ratio 0.04 [95% CI 0.01-0.07] vs 0.05 [95% CI 0.02-0.08], p=0.66) was similar between the two study groups (Fig 5b-c). In contrast, the abundance of neutrophils (median ratio 0.30 [95% CI 0.17-0.43] vs. 0.17 [95% CI 0.06-0.28], p=0.018) was higher in PLHIV than HIV-uninfected participants (Fig 5d).

**Figure 4.**
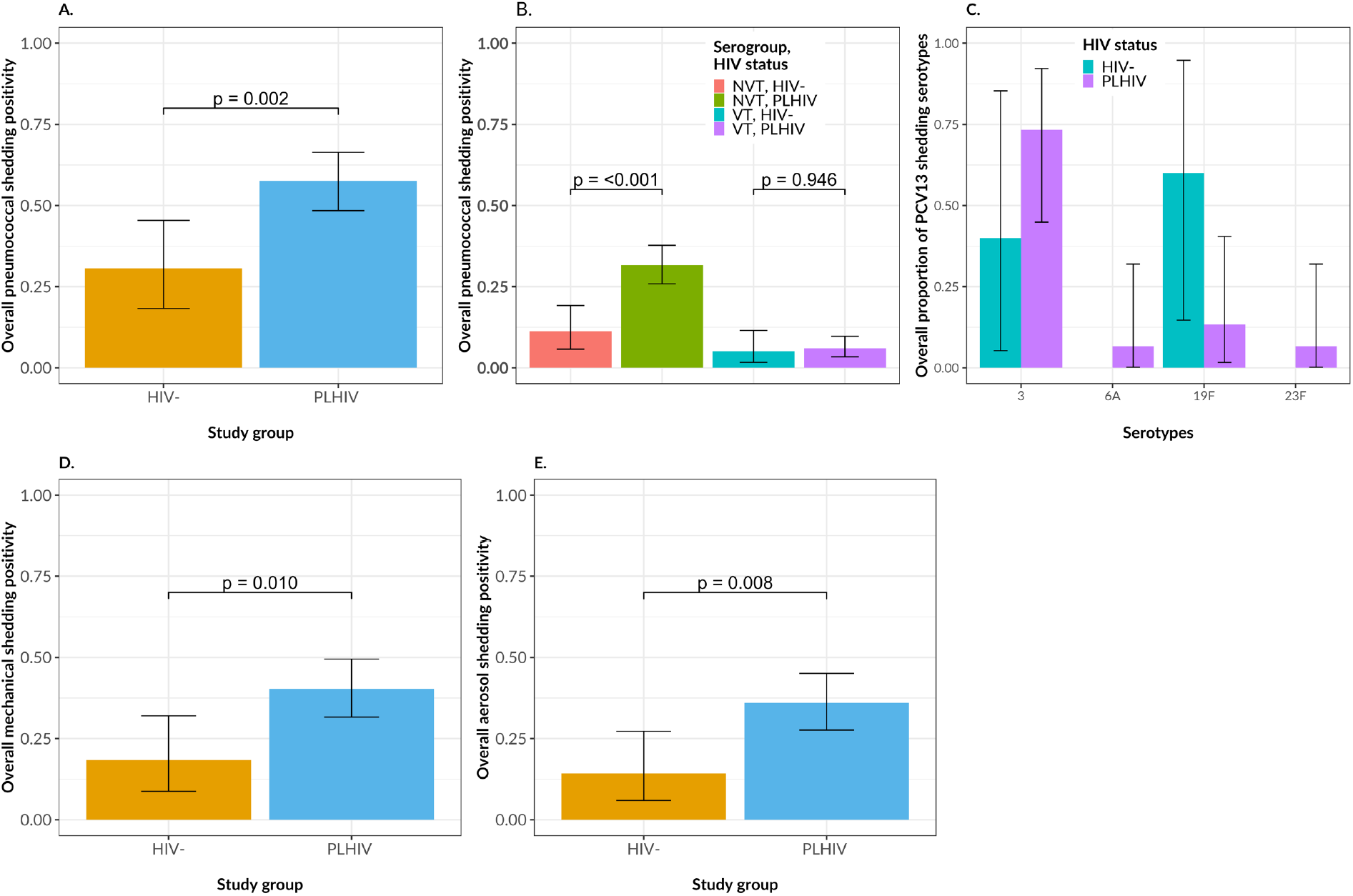
Pneumococcal shedding among PLHIV and HIV-uninfected adults. Pneumococcal shedding was defined as presence of viable pneumococci following microbiological culture of samples collected through nose poking (mechanical) and coughing or facemask sampling (aerosol). A) Overall pneumococcal shedding positivity stratified by HIV status. B). Overall pneumococcal shedding positivity stratified by HIV status and serogroup. C). Overall pneumococcal shedding positivity stratified by HIV status and PCV13 serotypes, the denominator for each serotype is the total PCV13 serotypes in each group. D). Pneumococcal mechanical shedding positivity stratified by HIV status E). Pneumococcal aerosol shedding positivity stratified by HIV status. The vertical bars represent median and horizontal lines represent confidence intervals (CI) (black lines). Data were analysed using Chi-square test (HIV-uninfected: n=23, PLHIV: n=61). Source data are provided as a Source Data file. ART, antiretroviral therapy

**Figure 5.**
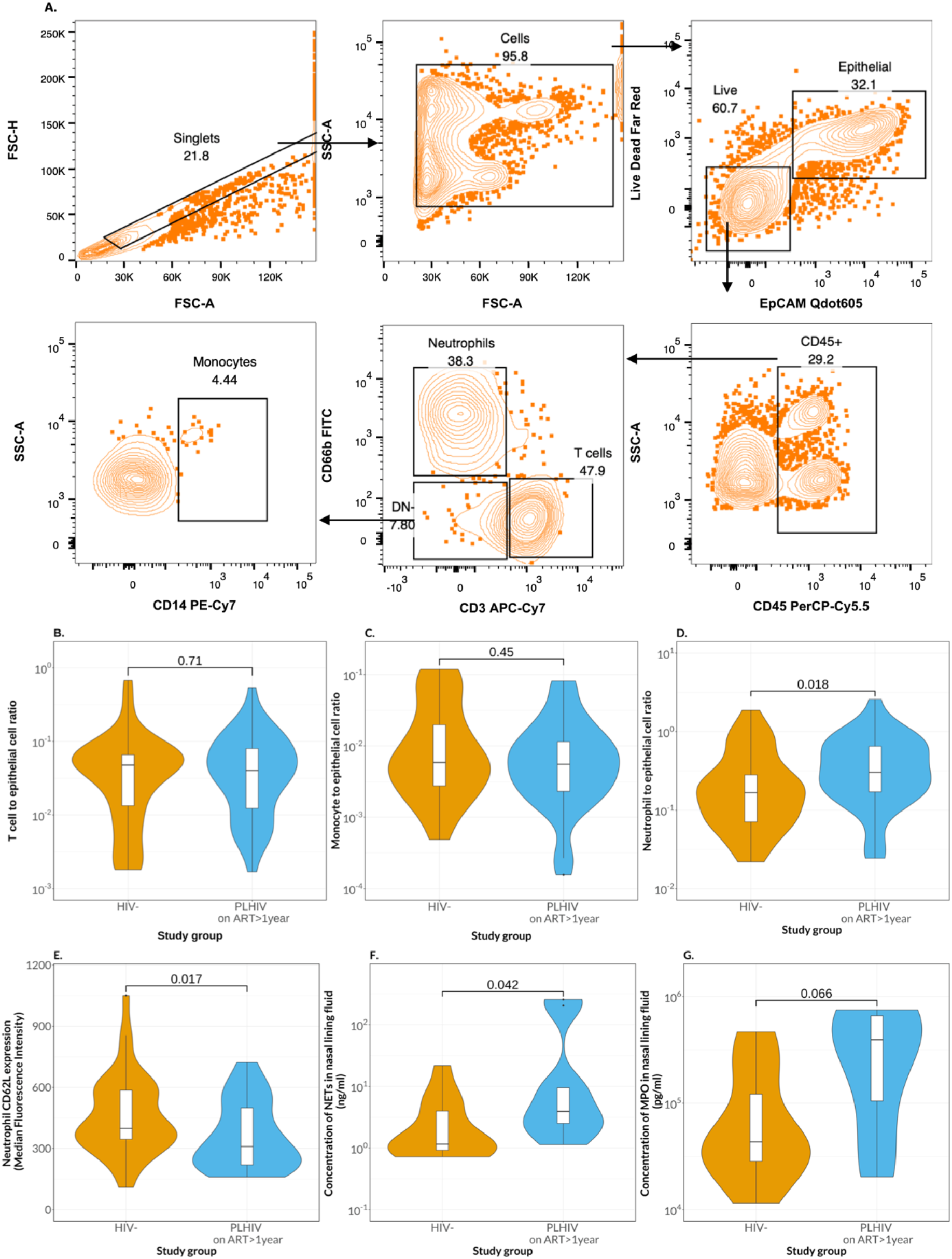
Nasal immune responses in adults with or without HIV. Immune cell profiling was done on nasal curette-collected mucosa cells using flow cytometry-based immunophenotyping, while concentration of NETs and MPO were measured in nasal lining fluid using ELISAs, in HIV-uninfected adults and PLHIV. Samples were collected weekly for 4 weeks in all participants. A) Representative flow cytometry plot from an HIV-uninfected participant showing a gating strategy for the identification of immune cells in the nasal mucosa. B-D) Plots showing abundance of immune cell subsets at all time points in the nasal mucosa of HIV-uninfected and PLHIV. Data was normalised as immune cell to epithelial cell ratio. E) Median fluorescence Intensity (MFI) of CD62L expression on neutrophils between HIV-uninfected and PLHIV at all time points. For all boxplots, box boundaries correspond to 25^th^ and 75^th^ percentiles; whiskers extend to a maximum of 1.5x the inter-quartile range, with values outside the box and whiskers being outliers. Data were analysed using Wilcoxon test (HIV-uninfected: n=10, PLHIV: n=9). Source data are provided as a Source Data file. ART, antiretroviral therapy

To get further understanding of the phenotype of neutrophils, we measured the expression of CD62L, which is down regulated during neutrophil activation ^34, 35^. Neutrophils from PLHIV had lower expression of CD62L than those from HIV-uninfected participants (Median fluorescent intensity (MFI) 310 [95% CI 264-356] vs. 399 [95% CI 345-453], p=0.017) (Fig 5e). Furthermore, PLHIV had higher concentrations of neutrophil pro-inflammatory markers in their nasal lining fluid including neutrophil extracellular traps (NETs) (3.9 ng/ml [95% CI 0-31.7] vs 1.2 ng/ml [95% CI 0-2.76], p=0.042) and myeloperoxidase (MPO) (393500 pg/ml [95% CI 311066-475934] vs 43425 pg/ml [95% CI 2008-84842], p=0.066), compared to HIV-uninfected participants (Fig 5f-g). Together, these findings show heightened inflammation in the nasal mucosa of PLHIV.

## Discussion

We assessed pneumococcal carriage dynamics in otherwise healthy PLHIV on ART for more than 1 year compared to HIV-uninfected adults. We found that PLHIV had high-density pneumococcal carriage and shed antimicrobial resistant *S. pneumoniae*. Moreover, PLHIV exhibited heightened neutrophil-associated inflammation in the nasal mucosa. These data highlight this vulnerable HIV-affected population as a potentially important reservoir for transmission of antimicrobial resistant pneumococci.

It is postulated that pneumococcal transmission is primarily through respiratory droplets ^36^ but direct contact through hands as a vehicle has also been implicated ^27, 28^. Consistent with this, we observed pneumococcal shedding in adults through nose poking and coughing. Unlike isolates obtained from the nose poking samples (nasopharyngeal niche), aerosol shed *S. pneumoniae* (likely from the oropharyngeal niche) from PLHIV were more frequently penicillin-resistant and multidrug-resistant. *Streptococcus mitis* and *Streptococcus oralis*, commensals of the human oropharynx, have been implicated as a potential source of antibiotic resistance genes for *S. pneumoniae* ^37, 38, 39, 40^, through horizontal gene transfer (HGT) of penicillin-binding protein (*pbp*) genes that drive emergence of β-lactam resistance ^41, 42^. An analysis of publicly available pneumococcal genomes has highlighted that *S. mitis* as a more frequent donor of genetically diverse *pbp* gene fragments to *S. pneumoniae* compared with *S. oralis*, with horizontal acquisition mostly confined to pneumococcal serotypes associated with prolonged nasopharyngeal carriage duration ^40^. These findings suggest that proximity between *S. pneumoniae* and other closely-related streptococci species harboring antibiotic resistance genes facilitates their transfer to pneumococci, and this could in part explain the predominance of penicillin-resistant pneumococci in the aerosol shed isolates.

On the other hand, it has been recently shown that there is an elevated risk of both the acquisition and persistent carriage of multi-drug resistant pneumococci following antimicrobial treatment ^43^. The mechanism of persistence of multi-drug resistant pneumococci was attributed to reduced within-host competition following antimicrobial treatment, indicating that in the absence of treatment, susceptible lineages outcompete resistant lineages ^43^. In Malawi, as part of standard HIV clinical management, PLHIV are put on daily co-trimoxazole prophylaxis ^17, 18^. Consistent with purifying selection and in agreement with previous studies ^44, 45^, there was higher prevalence of co-trimoxazole resistant carriage isolates in PLHIV than HIV-uninfected participants. Considering that co-trimoxazole is a broad-spectrum antimicrobial and its resistance is associated with penicillin resistance^45, 46^, it could differentially alter the microbiota in the oropharyngeal and nasopharyngeal compartments, resulting in stronger selection for multi-drug resistant pneumococci.

Consistent with their increased propensity for pneumococcal shedding, PLHIV exhibited neutrophil-driven nasal inflammation. Neutrophil factors including NETs and MPO contribute to clearance of pneumococcal infection ^47, 48^, but also promote excessive inflammation ^25, 49^. Host inflammation has been shown to drive pneumococcal shedding ^31, 32, 33 50^. In mice models, pneumococcal virulence factors, such as pneumolysin and pneumococcal surface protein K (PspK), induce upper respiratory tract (URT) inflammation resulting in pneumococcal shedding ^32, 33, 51^.

Furthermore, uncontrolled inflammation has been shown to promote pneumococcal persistence via nutrient provision arising from increased mucus secretion ^52 53, 54^, and through reduced mucocilliary clearance of pneumococci due to altered ciliary beating ^55^. Together, this suggests that uncontrolled nasal inflammation in PLHIV, likely due to impaired nasal immunity ^10, 56^ or driven by altered URT microbiota, perpetuates nasal inflammation leading to increased pneumococcal shedding, a surrogate of transmission potential.

Despite the strengths of this study, including a comprehensive assessment of carriage dynamics in a relevant population and setting, there are important limitations. First, the standard microbiological assay used in this study were biased towards isolating single isolates following culture, hence missing any potential multiple serotype carriage episodes or pneumococci carried at low densities. Second, we did not directly measure pneumococcal transmission. Although pneumococcal shedding is regarded as a surrogate for transmission potential ^27, 28^, it is not clear how often shed pneumococci result in successful transmission. However, this does not discount our suggestion that PLHIV on ART for more than 1 year contribute to transmission of multi-drug resistant pneumococci, which is of public health importance.

In conclusion, our study demonstrates that PLHIV on ART for more than 1 year could be an important reservoir for transmission of multi-drug resistant *S. pneumoniae*. This has potential to derail the success of the infant PCV programme and our fight against antimicrobial resistance in high HIV prevalence and high pneumococcal carriage settings such as Malawi, especially now that there is an HIV test and treat strategy ^57^. Considering that PCV was shown to be efficacious against recurrent IPD among PLHIV in Malawi^58^, consideration should be given to re-evaluating the provision of pneumococcal vaccination to this vulnerable adult population.

## Methods

### Study design and recruitment

We recruited asymptomatic PLHIV on ART for more than 1 year and HIV-uninfected adults in Blantyre, Malawi. Participants were recruited on day 3 after screening and then followed up on day 7, 14, 21 and 28 for the first month, and then every month for 5 months. All participants were recruited from the ART clinics and Voluntary Counselling and Testing (VCT) centres at Lighthouse-Queen Elizabeth Central Hospital and Gateway Health Centre in Blantyre. Briefly, participants were screened for pneumococcal carriage, using WHO recommendations for nasopharyngeal sampling and microbiological culture for pneumococcal detection ^59^. Inclusion criteria included confirmed pneumococcal carriage, in adults aged 18 to 45 years, living with a child aged under-5-years and providing written informed consent.

Exclusion criteria included receiving antibiotics in the previous 4 weeks (apart from co-trimoxazole prophylaxis as all PLHIV on ART receive daily co-trimoxazole prophylaxis as per national guidelines ^17^), hospitalization for pneumonia in the previous 4 weeks, having respiratory tract Kaposi’s sarcoma or a terminal illness (e.g., metastatic malignancy, terminal AIDS). All PLHIV were on standardised ART regimen according to Malawi Ministry of Health 2018 Clinical Management of HIV guidelines ^60^.

The study was conducted in accordance with good clinical practice (GCP) guidelines and the Declaration of Helsinki. Ethical approval was obtained from the College of Medicine Research Ethics Committee (COMREC) (P.11/18/2532) and Liverpool School of Tropical Medicine Research Ethics Committee (LSTMREC) (19-033).

### Sample collection

Nasopharyngeal swabs were collected at all time points, and a peripheral blood sample was collected at recruitment (day 3) from PLHIV on ART for measuring CD4 count. In addition, respiratory secretions were collected at day 3, 21, and 28, using a modified polyvinyl acetate (PVA) facemask device ^61, 62, 63^, direct coughing onto an agar plate (coughing), and inserting the participants clean index finger into the front of the nose (nose poking). Further details of these methods are discussed in the supplementary section.

Nasal cells were collected weekly (5 visits) by scraping the inferior turbinate using nasal curette (Rhino-probe, Arlington Scientific, UK) as previously described ^64^. Per individual and timepoint, two curettes were collected from each nostril. Briefly, the four curette samples were immediately placed in a 15-ml falcon tube (Corning) containing 8-ml of precooled transport media (RPMI-1640 [Thermo Fisher Scientific Inc.) supplemented with 10% Fetal Bovine Saline [FBS], antibiotic/antimycotic [Sigma, USA], L-glutamine, non-essential amino acids, HERPES buffer and sodium pyruvate (all from Gibco, USA)).

Nasal-lining fluid was collected weekly (5 visits) using a non-invasive method that uses a synthetic absorptive matrix (SAM) strip (Hunts Development Ltd, UK). Briefly, the nasosorption strip was advanced up the lumen of the nasal cavity and held inside the nostril for up to three minutes until soaked. The strip was then removed and placed in a microcentrifuge tube for storage at −80°C until processing.

### Microbiological culture and density quantification

Standard microbiologic culture was used to determine the presence of *S. pneumoniae* from the nasal swab, nose poke swab, cough onto an agar plate and modified PVA facemask^61, 62, 63^. For modified PVA facemask processing, four PVA strips were removed from the mask using sterile technique and then dissolved in 10ml Todd Hewitt broth with 5% yeast (THY) and then plated. We identified *S. pneumoniae* by their morphology and optochin sensitivity on a gentamicin-sheep blood agar plate (SBG; 5% sheep blood agar, 5μL gentamicin/mL) after an overnight incubation at 37 °C in 5% CO2. The bile solubility test was used on isolates with no or intermediate (zone diameter < 14mm) optochin susceptibility. Plates showing no *S. pneumoniae* growth were incubated for a further 24 hours before being reported as negative. A single colony of confirmed pneumococcus was selected and grown on a new SBG plate as before. Growth from this second plate was used for serotyping by latex agglutination (ImmuLexTM 23-valent Pneumotest; Statens Serum Institute, Denmark). This kit allows for differential identification of each PCV13 vaccine serotypes (VT) but not for differential identification of PCV13 non-vaccine serotypes (NVT); NVT and non-typeable isolates were, therefore, reported as NVT. Pneumococcal density was quantified using microbiological culture serial dilutions ^65^ on a gentamicin-sheep blood agar plate (SBG; 5% sheep blood agar, 5μL gentamicin/mL) and results reported as colony forming units per millilitre (CFU/ml).

### AMR profiling

Pure pneumococcal isolates were cultured on sheep blood agar without antibiotics (SBA), using an overnight incubation at 37 °C in 5% CO2. Growth from the SBA were emulsified in normal saline to match 0.5 McFarland turbidity standard and plated on sheep blood Muller Hinton agar plate without antibiotics (MHB) using a sterile cotton swab. The antimicrobial susceptibility of pneumococcal isolates was assessed by the disk diffusion method (Oxoid, USA) for beta lactams antibiotic (oxacillin 1μg), macrolide-lincosamide-streptogramin (MLS) antibiotic (erythromycin 15μg), tetracycline antibiotic (tetracycline 30μg) and Trimethoprim/sulfamethoxazole (co-trimoxazole 1.25–23.75μg).

Beta lactams antibiotic susceptibility was confirmed by benzylpenicillin E-test minimum inhibitory concentration (MIC). E-test MICs were determined by suspending colonies from an overnight culture (incubation at 37 °C in 5% CO2) on SBA in saline, and the organism density was matched to a 0.5 McFarland turbidity standard. Using a sterile cotton swab, the organism suspension was plated onto MHB. E-test strips were applied to the surface of the plates according to the manufacturer’s recommendations. Agar plates were incubated overnight at 37 °C in 5% CO2. The MIC was defined as the lowest concentration of drug where the zone edge intersected the E-test strip.

Interpretation of results followed European Committee on Antimicrobial Susceptibility Testing (EUCAST) guidelines ^66^. *Streptococcus pneumoniae* ATCC 49619 was used as quality control. Multidrug resistance (MDR) was defined as non-susceptibility to agents in three or more above chemical classes of antibiotic.

### Flow cytometry analysis

For immunophenotyping, nasal cells were dislodged from curettes by repeated pipetting and stained with far-red fluorescent amine reactive dye (34973A, 1:1000) from Invitrogen, and incubated for 15 minutes followed by the addition of a cocktail of BV605 anti-human CD326 (9C4, 324224, 1:50), PerCp-Cy5.5 anti-human CD45 (HI30, 304028, 1:40), FITC anti-human CD66b (G10F5, 305104, 1:40), PE-Cy7 anti-human CD14 (M5E2, 301814, 1:40), APC-Cy7 anti-human CD3 (SK7, 344818, 1:40) and PE-eFlour® 610 anti-human CD62L (DREG-56, 61062942, 1:40), all from Bio legend (UK). Samples were acquired on LSR FORTESSA flow cytometer equipped with FACSDIVA version 8.0.1 (BD Biosciences, UK) and analysed using Flowjo Version 10.8.1 (BD Biosciences, USA). Abundance of immune cells was normalised as immune cell to epithelial cell ratio as previously reported ^67^. All flow cytometry data are available.

### Neutrophil-extracellular traps (NETs) enzyme linked-immunosorbent assay

Nasosorption samples were eluted from stored nasosorption filters (Mucosal Diagnostics, Hunt Development (UK) Ltd, Midhurst, UK) using 200μL of elution buffer (Millipore) by centrifugation at 1500g for 10 minutes as previously described ^68^. The eluate was cleared by further centrifugation at 1595g for 10 minutes. The eluate was then stored at −80°C until the day of the assay. The detection of NETs was performed by combining two commercial ELISA kits; ROCHE cell death ELISA kit (Sigma Aldrich, 11920685001) and Human Elastase ELISA Kit (Hycult®Biotech, HK319). In-house developed PMA NETs were serially diluted (1:2) and used as standards, while nasosorption samples were diluted (1:4) using 1X dilution buffer from the Elastase ELISA Kit (Hycult®Biotech, HK319).

### Myeloperoxidase (MPO) enzyme linked-immunosorbent assay

Myeloperoxidase (MPO) enzyme linked immunosorbent assay was performed on eluted −80°C frozen nasal lining fluid samples as described in the manufacturer’s user guide (Invitrogen, UK, BMS2038INST).

### Statistical analysis

Descriptive statistics are reported as numbers and proportions for categorical variables, and median and confidence interval for continuous data. Categorical variables were compared using χ2 test, and continuous variables were compared using Mann-Whitney test. We fitted a univariate and multivariable regression model, accounting for multiple sampling (generalized linear mixed model), covariables used for each model are detailed in the supplementary section. All data analyses were done in RStudio (Version 4.1.3, R Development Core Team, Vienna, Austria). Figures and tables were produced in R (v4.1.3), RStudio (v2022.02. 2+485.pro2 Prairie Trillium - April 27, 2022), ggplot2 (v3.3.5), lme4 (v1.1-28), and gtsummary (v1.5.2)^69^. Statistical significance level was reported at a p value of <0.05.

## Data Availability

All data produced in the present study are available upon reasonable request to the authors

## Data availability

All data supporting these findings are available upon reasonable request.

## Acknowledgements

The authors thank all study participants, and the staff of the Queen Elizabeth Central Hospital (QECH) Lighthouse and Gateway Clinic, for their support and co-operation during the study. This work was supported by the National Institute for Health Research (NIHR) [16/136/46]. KCJ is supported by an MRC African Research Leader award [MR/T008822/1]. RSH is a NIHR senior investigator. The views expressed in this publication are those of the authors and not necessarily those of the NIHR or the UK government. A Wellcome Strategic award number 206545/Z/17/Z supports MLW. The funders were not involved in the design of the study; in the collection, analysis, and interpretation of the data; and in writing the manuscript. The findings and conclusions in this report are those of the authors and do not necessarily represent the official position of the funders.

## Author contributions

The author contributions were as follows: Methodology: K.C.J., R.S.H. and N.F. Investigation: L.S., J.P., N.M., N.K., T.N., A.K., M.K., and G.S. Data analysis: K.C.J., L.S., and J.P. Interpretation: K.C.J., L.S., J.P., R.S.H., B.K.A, T.D.S., and N.F. Data curation: L.S. and J.P. Project administration: N.M., N.K. and K.C.J. Sample collection: A.K., M.K., and G.S. Writing: L.S., J.P., and K.C.J. Conceptualisation and supervision: K.C.J., R.S.H., B.K.A, and N.F. All authors read and approved the final manuscript.

## Corresponding author

Correspondence to Kondwani C. Jambo

