## Supplementary Materials for "Frequent shedding of multi-drug resistant pneumococci among adults living with HIV on suppressive antiretroviral therapy in Malawi"

### Methods

*Collection of shedding samples*

To collect respiratory secretions participants were asked to wear the modified PVA facemask containing four 1×9 cm 3D printed polyvinyl-alcohol (PVA) sampling matrix strips placed horizontally across the inside of the mask for 15 minutes [1, 2, 3], and they were asked to cough every 5 minutes during the 15 minutes period and the clinical nurses kept the participant in a conversation during the period. To collect nose poking samples participants were asked to pick their nose using an index finger and using a wooden cotton swab clinical nurses swabbed the participant index finger and placed the swab in transport media. To collect the secretion from a cough, participants were asked to cough directly onto a gentamycin-sheep blood agar plate (SBG; 5% sheep blood agar, 5µL gentamicin/mL). All the samples were sent to the laboratory within an hour of collection.

*Neutrophil-extracellular traps (NETs) enzyme linked-immunosorbent assay*

50uL of standards and nasosorption samples were added to appropriate wells of the pre-coated Elastase ELISA plates and incubated for two hours at room temperature with agitation (350rpm, flat shaker). After incubation, the plate was washed four times with 1X wash buffer (from the Elastase ELISA kit). Anti-DNA-POD (From Cell Death Detection ELISA$^{PLUS}$, 10x kit, 11920685001) was added followed by incubation at room temperature for 2 hours with agitation (350rpm, flat shaker). The plate was washed five times with wash buffer (Hycult®Biotech, HK319) and 100µL of TMB substrate was added. The plate was incubated for 5 minutes at room temperature with agitation (350 rpm, flat shaker) before 100ul of stop solution (Elastase ELISA Kit) was added. The endpoint absorbance readings were measured on a microplate reader (BioTek ELx808, UK) at 450 nm, and a standard curve was plotted using 4-parameter or hyperbola curve, and the concentration of NETs was calculated using the standard curve.

*Myeloperoxidase (MPO) enzyme linked-immunosorbent assay*

The instant Human MPO ELISA kit is pre-coated with capture antibody and contains streptavidin HRP and biotin conjugate in the wells. Moreover, assay standards were prediluted and added to the standard well strips in a lyophilised form by the kit manufacturer. In Brief, nasosorption samples were diluted (1:100) in sample diluent. Then 100uL of pre-diluted samples were added to precoated wells containing all the necessary reagents. The plate was incubated on a microplate shaker at room temperature for 3 hours. After incubation, the plate was washed four times in 1X wash buffer (phosphate buffered saline with 1% tween ™ 20), and 100uL of TMB substrate solution (tetramethyl-benzydine) was added. The plate was incubated for 10 minutes at room temperature, before addition of 100ul of Stop solution (1M phosphoric acid). The optical density (OD) of each well was read at 450 nm in a microplate reader (BioTek ELx808, UK). The average OD values for each set of duplicate standards and samples was calculated. A standard curve was plotted sing a 5-parameter curve fit and the concentration of MPO was calculated.

*Statistical analysis*

Pneumococcal colonisation episode was defined by either the first pneumococci carriage at study screening or re-acquisition of a pneumococci after pneumococcal clearance. Pneumococcal clearance was referred to as the detection of negative cultures for any serotype at two consecutive sampling points as previously described [4, 5, 6]. Screening carriage was assumed to occur 1 week before and for episodes which commenced from screening and terminating before week 4, acquisition and clearance of the serotype was assumed to occur at the mid-point between consecutive sampling points. From week 4 to week 20 where samples were collected bi-monthly, acquisition and clearance of serotypes was assumed to occur 1 week before and after detection to prevent overestimation of the carriage duration.

*Definition of variables for analysis:* Pneumococcal carriage duration was estimated not as a duration per isolate but as the duration of carriage state in an individual, with two consecutive negative carriage results considered as clearance as previously reported [4, 5]. Pneumococcal carriage duration of less than 50 days was considered short duration, while carriage density of less than 2010CFU/ml was considered low density carriage, which represents values below the 25$^{th}$ percentile (Fig. S2a-b). While CD4 count of less than 435Cells/µL was considered low CD4 count, and ART duration of less 3.5years was considered short ART duration, which represents values below the 25$^{th}$ percentile (Fig. S2c-d). Possession index was considered a surrogate of social economic status, with a higher value representing higher social economic status. Season was defined as Hot wet (November and April) and Cool dry (May and October) (according to Malawi Meteorological Services).

*Covariables for logistic regression analysis:* To identify factors influencing pneumococcal carriage prevalence the following covariables were included age, gender, number of children, possession index, season, and HIV status. To identify factors influencing pneumococcal carriage duration the following covariables were included age, gender, median carriage density per episode, possession index, season, and HIV status. To identify factors influencing pneumococcal carriage density the following covariables were included age, gender, possession index, season, HIV status. All covariables were chosen priori based on their potential to influence pneumococcal carriage dynamics.

Supplementary Tables

Table S1

Supplementary Table 1. Overall Baseline characteristics

| Characteristic | **HIV-**, N = 54[1] | **PLHIV on ART >1yr**, N = 90[1] | **p-value**[2] |
|---|---|---|---|
| **Sex** | | | 0.5 |
| Female | 34 (63%) | 62 (69%) | |
| Male | 20 (37%) | 28 (31%) | |
| **Age group** | | | **0.059** |
| 18-25 | 20 (37%) | 19 (21%) | |
| 26-35 | 21 (39%) | 35 (39%) | |
| 36-45 | 13 (24%) | 36 (40%) | |
| **Number of children** | | | **0.028** |
| Median (IQR) | 1.00 (1.00, 2.00) | 1.00 (1.00, 1.00) | |
| **Socioeconomic status** | | | 0.11 |
| Median (IQR) | 7.00 (3.00, 8.00) | 5.00 (4.00, 6.00) | |
| **CD4 count (Cells/µl)** | | | **<0.001** |
| Median (IQR) | 786 (673, 945) | 560 (349, 773) | |
| **ART duration (Years)** | | | |
| Median (IQR) | NA (NA, NA) | 5.5 (2.8, 10.1) | |
| **HIV viral load (Copies/ml)**[3] | | | |
| Median (IQR) | NA (NA, NA) | 39 (39, 25,150) | |

[1] n (%)

[2] Pearson's Chi-squared test; Wilcoxon rank sum test

[3] i.e. Only 13 PLHIV on ART>1yr had a detectable Viral load

Table S2a

| Supplementary Table 2a. NVT carriage vs NVT no carriage | | | | | | | |
|---|---|---|---|---|---|---|---|
| | | **Univariate** | | | **Multivariable** | | |
| **Characteristic** | **N** | **OR[1]** | **95% CI[1]** | **p-value** | **OR[1]** | **95% CI[1]** | **p-value** |
| **HIV status** | 900 | | | | | | |
| HIV- | | — | — | | — | — | |
| PLHIV on ART >1yr | | 1.53 | 1.17, 2.00 | **0.002** | 1.43 | 1.07, 1.90 | **0.015** |
| **Sex** | 900 | | | | | | |
| Female | | — | — | | — | — | |
| Male | | 0.93 | 0.69, 1.25 | 0.6 | 0.97 | 0.71, 1.33 | 0.9 |
| **Age group (Years)** | 900 | | | | | | |
| 18-25 | | — | — | | — | — | |
| 26-35 | | 0.81 | 0.58, 1.14 | 0.2 | 0.73 | 0.51, 1.04 | 0.080 |
| 36-45 | | 1.28 | 0.90, 1.81 | 0.2 | 1.08 | 0.74, 1.57 | 0.7 |
| **Season** | 898 | | | | | | |
| Cooldry | | — | — | | — | — | |
| Hotwet | | 0.93 | 0.71, 1.22 | 0.6 | 0.93 | 0.71, 1.23 | 0.6 |
| **Socioeconomic status** | 900 | | | | | | |
| Medium/high ses (> 3) | | — | — | | — | — | |
| Low ses (<= 3) | | 1.23 | 0.93, 1.61 | 0.14 | 1.19 | 0.90, 1.59 | 0.2 |

[1] OR = Odds Ratio, CI = Confidence Interval

Table S2b

| Supplementary Table 2b. VT carriage vs VT no carriage | | | | | | | |
|---|---|---|---|---|---|---|---|
| | | Univariate | | | Multivariable | | |
| **Characteristic** | **N** | **OR**[1] | **95% CI**[1] | **p-value** | **OR**[1] | **95% CI**[1] | **p-value** |
| **HIV status** | 900 | | | | | | |
| HIV- | | — | — | | — | — | |
| PLHIV on ART >1yr | | 1.08 | 0.19, 6.03 | >0.9 | 1.02 | 0.71, 1.46 | >0.9 |
| **Sex** | 900 | | | | | | |
| Female | | — | — | | — | — | |
| Male | | 0.92 | 0.14, 6.23 | >0.9 | 1.02 | 0.69, 1.52 | >0.9 |
| **Age group (Years)** | 900 | | | | | | |
| 18-25 | | — | — | | — | — | |
| 26-35 | | 1.60 | 0.16, 15.9 | 0.7 | 1.51 | 0.95, 2.40 | 0.084 |
| 36-45 | | 1.58 | 0.15, 16.9 | 0.7 | 1.45 | 0.88, 2.39 | 0.14 |
| **Season** | 898 | | | | | | |
| Cooldry | | — | — | | — | — | |
| Hotwet | | 0.74 | 0.53, 1.04 | 0.084 | 0.77 | 0.55, 1.08 | 0.13 |
| **Socioeconomic status** | 900 | | | | | | |
| Medium/high ses (> 3) | | — | — | | — | — | |
| Low ses (<= 3) | | 1.05 | 0.19, 5.90 | >0.9 | 1.01 | 0.71, 1.44 | >0.9 |

[1] OR = Odds Ratio, CI = Confidence Interval

Table S3

Supplementary Table 5. Pneumococcal shedding isolates antibiogram among HIV uninfected adults

| Characteristic | Aerosol shedding vs Nasopharyngeal carriage | | | Mechanical shedding vs Nasopharyngeal carriage | | |
|---|---|---|---|---|---|---|
| | **Aerosol shedding**, N = 7[1] | **Nasopharyngeal carriage**, N = 7[1] | **p-value**[2] | **Mechanical shedding**, N = 9[1] | **Nasopharyngeal carriage**, N = 9[1] | **p-value**[2] |
| **Tetracycline** | 6 (86%) | 5 (71%) | >0.9 | 6 (67%) | 6 (67%) | >0.9 |
| **Erythromycin** | 6 (86%) | 4 (57%) | 0.6 | 6 (67%) | 6 (67%) | >0.9 |
| **Cotrimoxazole** | 6 (86%) | 5 (71%) | >0.9 | 9 (100%) | 8 (89%) | >0.9 |
| **Benzylpenicillin**[3] | 4 (57%) | 5 (71%) | >0.9 | 3 (33%) | 3 (33%) | >0.9 |
| **MDR** | 6 (86%) | 5 (71%) | >0.9 | 6 (67%) | 6 (67%) | >0.9 |

[1] n (%)

[2] Fisher's exact test

[3] i.e. Minimum inhibitory concentration

Supplementary Figures

Figure S1

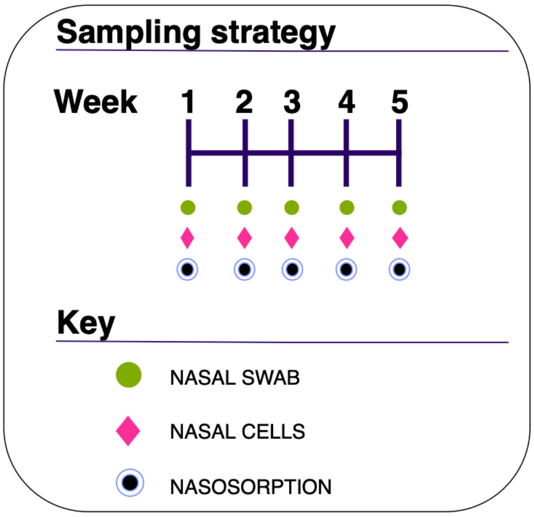


**Figure S1: Figure legend.** Sampling schedule showing different samples collected from all study participants at baseline and their follow-up. Nasopharyngeal swab, Nasosorption and nasal cells were collected every week for 5 weeks.

### Figure S2

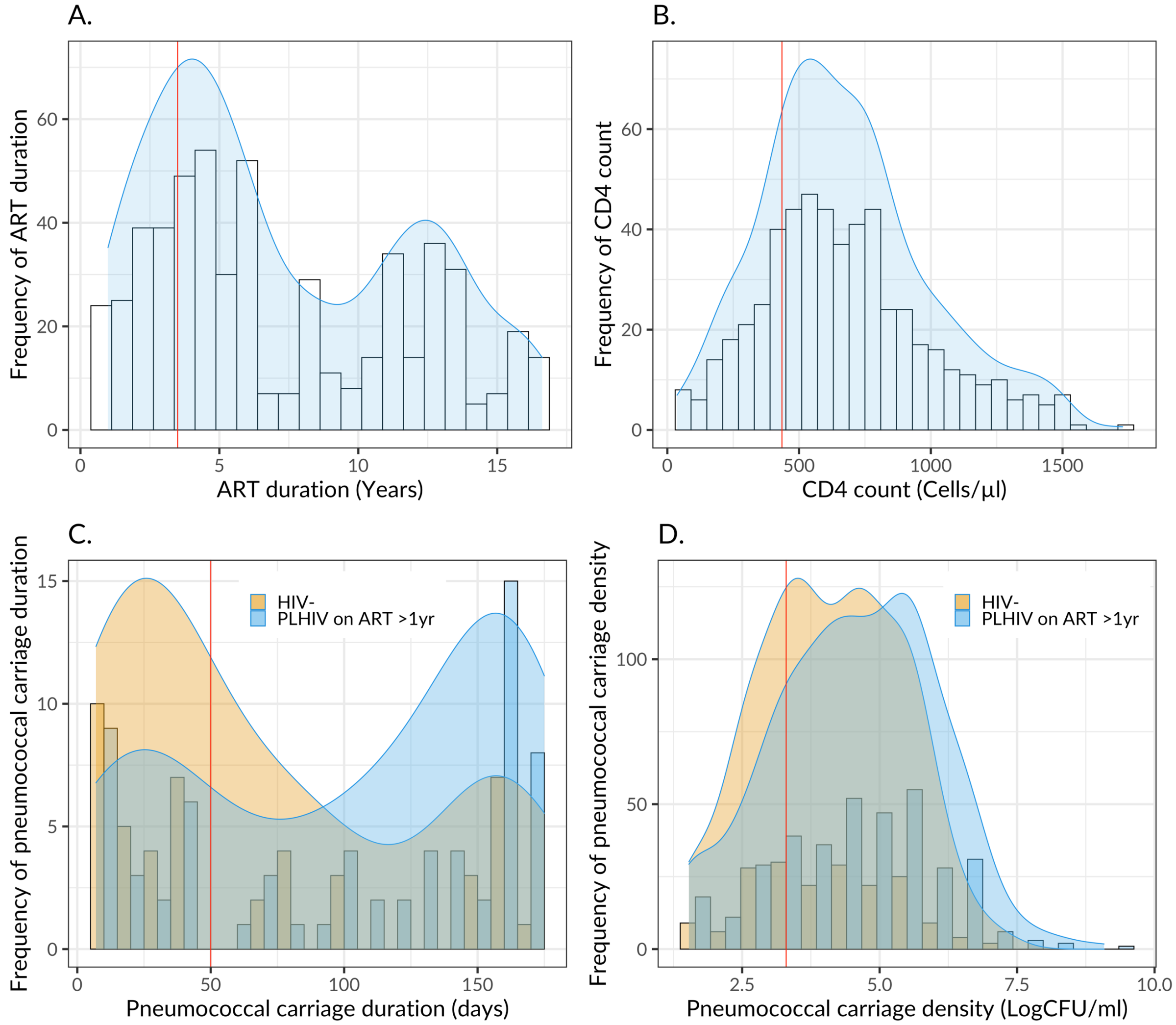


**Figure S2: Figure legend.** a) Histogram showing distribution of ART duration in year among PLHIV. b) Histogram showing distribution of CD4 count (Cells/□l) among PLHIV. c) Histogram showing distribution of pneumococcal carriage duration stratified by HIV status. d). Histogram showing distribution of pneumococcal carriage density stratified by HIV status. The red line indicates the 25th quantile used in the multivariate analysis.
